# Detection of multidrug resistant organisms of concern including *S. maltophilia* and *B. cepacia* at a referral hospital in Kenya

**DOI:** 10.1101/2023.06.12.23291242

**Authors:** Racheal Kimani, Patrick Maina, Moses Kamita, David Mbogo, Winnie Mutai, Charchil Ayodo, Essuman Suliman, Bernard N. Kanoi, Jesse Gitaka

## Abstract

It is important to monitor of antibiotic susceptibility patterns of bacteria in clinical settings periodically to ascertain the current trends as well as re-establish empirical therapy. This study aimed to determine bacterial contaminants and their antimicrobial susceptibility patterns from medical equipment, inanimate surfaces and clinical samples isolated from Thika Level V Hospital (TLVH). Three hundred and five samples were collected and comprised of urine, pus swabs, catheter swabs, stool and environmental samples. Bacterial identification and antimicrobial susceptibility testing were done on VITEK 2 and disc diffusion respectively.

Coagulase negative *Staphylococci* (28 /160, 17.5%) were the most isolated species from patients followed by *E. coli* (22 /160 13.8%) and *S. aureus* (22/160, 13.8%). The bed rail was the most contaminated surface with *S. aureus* at (6/42)14.2%. The clinical sample that yielded the highest number of pathogens was pus (92/160). Trauma patients had the largest proportion of isolates (67/160, 41.8%). Bacteria recovered from this study demonstrated high levels of resistance especially enteric bacteria. Extended Spectrum Beta Lactamase phenotype was noted in 29/44 (65.9%) enteric isolates. Although ESBL genetic confirmatory studies are needed, this study shows that there is an urgent need for actions that mitigate the spread of antibiotics resistant bacteria.

## Introduction

The emergence of multi-drug resistant (MDR) strains in health care facilities poses a threat in the management of infections in developing countries (1). Approximately 700,000 people die annually from antimicrobial resistant (AMR) infections globally (2), with a projected 10 million deaths by 2050(3). In Kenya, high levels of hospital-acquired infections (HAI’s) have been reported signifying the need for reinforced infection prevention control strategies (4) (5) (6). Multidrug-resistant bacterial contaminants have been reported in hospital wards in Kenya (7) albeit in a small number of hospitals.

At present, β-lactam drugs are a key factor in the treatment of bacterial infections worldwide and account for almost 65% of antibiotic usage (1)drugs have been classified into six main groups based on the chemical structure of the β-lactam ring and includes penicillin, cephalosporins, cephamycin, carbapenems, monobactams, and β-lactamase inhibitors. Nevertheless, it is unfortunate that, in recent years, resistance to this important class of antibiotics has been shown to increase globally (8).

Bacterial infections are a common complication in patients admitted in hospitals for other reasons other than the infections. Those who acquire HAIs are at an increased risk of longer hospital stay, which increases medical care cost (9). Nevertheless, there are patients also seeking medical attention due to community-acquired infections (CAIs).

Incidentally, both CAI’s and HAI’s causative agents have registered increasing resistance to antibiotics which has reached an alarming level for some organisms (10). The hospital environment is a source multidrug resistant organisms (MDROs) especially when environmental disinfection, contact precautions and hand hygiene are not routinely employed (11). High-risk areas are hard surfaces like beds, handles and grab bars. Transmission of pathogens to patients is thought to be through healthcare workers, visitors, or asymptomatic carriers (12). Enhanced cleaning has been emphasized to overcome transmission of MDROs. However, a study indicated that only 47% of these surfaces are cleaned (13). Pathogens reported to persist in healthcare environments include *Clostridium difficile* spores, vancomycin-resistant Enterococcus (VRE), methicillin-resistant *Staphylococcus aureus* (MRSA) and *Acinetobacter baumannii* (14)

CAIs, resistance is attributable to the widespread use of broad-spectrum antibiotics, the sale of antibiotics over the counter, self-treatment with antibiotics, the inappropriate use of antibiotics and a lack of compliance with treatment (15,16)

As for HAIs, causative bacteria resistance to antimicrobials is mainly due to the use of broad-spectrum antibiotics to control them. Among the microorganisms usually implicated in HAIs include *Pseudomonas aeruginosa, Escherichia coli, Staphylococcus aureus, Klebsiella species*, and coagulase-negative *Staphylococci*. Some of the factors that contribute to development of HAIs include hospital design factors (e.g., ventilation and adequate number of handwash basins)(17), longer hospital stays, gender, surgery since admission, intubation, mechanical ventilation, age of the patient, type of hospital, and urinary catheter and hygienic practices (18).

Here we sampled bacteria from patients as well as environment contaminating bacteria. We further determined their antimicrobial susceptibility patterns.

## Material and Methods

### Study population and samples collection

We collected 305 hospital samples at Thika Level V Hospital between March–November 2021. 200 were from patients and 105 form high contact surfaces. Patient sociodemographic, and clinical data (diagnosis and treatment received) were filled in pathological data forms. Urine, pus swabs, stool, and indwelling catheter swabs were aseptically collected from patients. The age bracket of our participants was 17 years to 96 years. Environmental swab samples from high contact surfaces including the bed rail, intravenous fluid pole, cabinets, medicine tray, monitors, nurses working stations, doorknobs and sphygmomanometer were also collected. Sampling was done by use of a moist sterile swab passed over the surfaces of inanimate objects and equipment. The patients sampled included those from the renal unit, the ICU, the male medical and surgical wards (male and female). A proportion of the renal unit patients were not admitted in the hospital. However, we didn’t establish if our patients had community acquired infections at the time of admission for surgery. Just a small proportion in the medical ward were primarily hospitalised due to infections (those with cellulitis, gangrene, and diabetic foot).

### Sample processing

The samples were transported under chilled conditions to the Mount Kenya University, Centre for Research, and Infectious Diseases Laboratory, for processing where they were cultured on selected media plates. Confirmatory identification was performed on VITEK 2 using gram negative (GN) and positive (GP) identification cards (ID). Quality control organisms included *Staphylococcus saprophyticus* (ATCC BAA-750) for GP ID Card and *Enterobacter cloacae* (ATCC 700323) for the GN ID Card.

### Antimicrobial susceptibility testing for Gram-negative bacteria

Antimicrobial susceptibility tests were performed by the standard Kirby–Bauer disk diffusion method on the Mueller–Hinton agar media (Thermo Scientific™ Oxoid™) using commercially available antibiotic disks (Oxoid™, UK). The diameter of inhibition zone was measured for each antibiotic disk, and the results were defined in accordance with the CLSI guidelines (CLSI, 2020). The antibiotics used included cefuroxime (30μg), ceftriaxone (30μg), cefotaxime (30μg), piperacillin (30μg), sulphamethoxazole (25μg), amoxicillin-clavulanate (10μg), gentamycin (10μg), aztreonam (30μg), ceftazidime (30μg), ciprofloxacin (5μg), cefoxitin (30μg), cefepime (30μg), minocycline (30μg), levofloxacin (5μg), amikacin (30μg), piptazobactam (110μg), imipenem (10μg), meropenem (10μg), tetracycline (30μg), nalidixic acid (10μg) and ampicillin (10μg). Quality control testing was achieved using ATCC microorganisms *E. coli* 25922 and *K. pneumoniae* 700603. Isolates that were resistant to cefotaxime, ceftriaxone, ceftazidime and aztreonam were presumed to be ESBL producers.

### Ethical considerations

This study was approved by the Research and Ethics committee of Mount Kenya University (MKU/ERC/1963) and licensed by National Commission for Science, Technology, and Innovation (NACOSTI) (NACOSTI/P/18/33304/25986). Before enrolment, informed consent was obtained from all participants or their legal guardians. Anonymized patient’s demographic characteristics were recorded on pathological investigation forms, which were filed and kept under lock and key, where they remained confidential.

### Data Analysis

The socio-demographic, clinical diagnosis, treatment received, and microbiological data were collectively documented for each patient on Microsoft Excel sheet. The environmental data were also documented accordingly. Kruskal–Wallis nonparametric test was used to test the differences in resistance between species using the SPSS statistical package version 26. Discrete data was described in means, and categorical data in frequencies and the data was presented in tables and figures.

## Results

### Patients baseline information

The mean age of participants was 46.11 years, and the range was 17 years to 96 years. Majority of positive cultures were from age group 27-36 (39/160; 24.3%) and 37-46 (34/160, 21.2%) years old as seen in **Table 1**. Patient’s presentations ranged from those with decubitus ulcers, burns, peritonitis, trauma, septic wounds, gangrene, diabetic foot, end stage renal disease, cellulitis, and congestive cardiac failure. Among the antibiotics received by patients were mainly ceftriaxone, metronidazole and flucloxacillin.

**Table 1:**
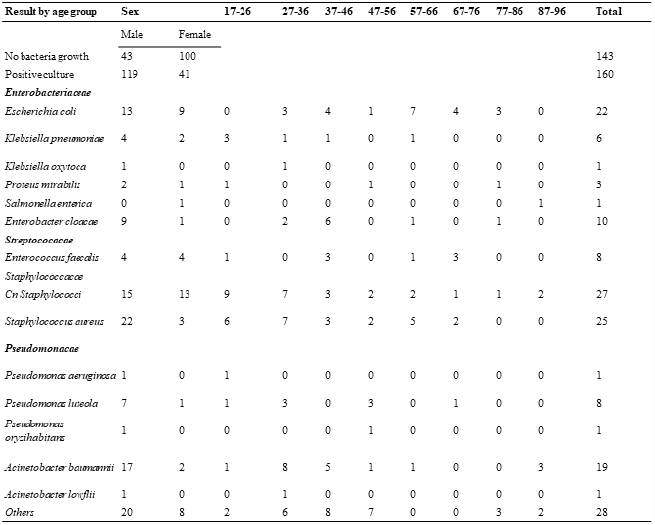
Demographic information of the participants and distribution of pathogens by age group

### Bacterial culture

Out of 305 culture positive plates (excluding mixed growth and fungal growth), 202 (66.2%) were culture positive; 160 from patients and 42 from hospital surfaces and equipment.

The highest prevalence of isolates was found from surgical ward with 72/202 (35.6%) isolates, followed by intensive care unit with 49/202 (24.2%), renal unit with 45/202 (22.2%) isolates and the medical ward with 36/202 (17.8%) isolates **(Figure 1a)**. Gram-negative isolates (55.4%, 112/202) were more than the gram-positive isolates (90/202, 44.6%). **Figure 1b and Figure 1c** shows all isolates identified for this study. Gram-positive isolates included CONS, *S. aureus, Enterococci* spp amongst others. Gram-negative species were mainly enteric bacteria, *Acinetobacter* spp, *Pseudomonas* spp, amongst others.

**Figure 1.**
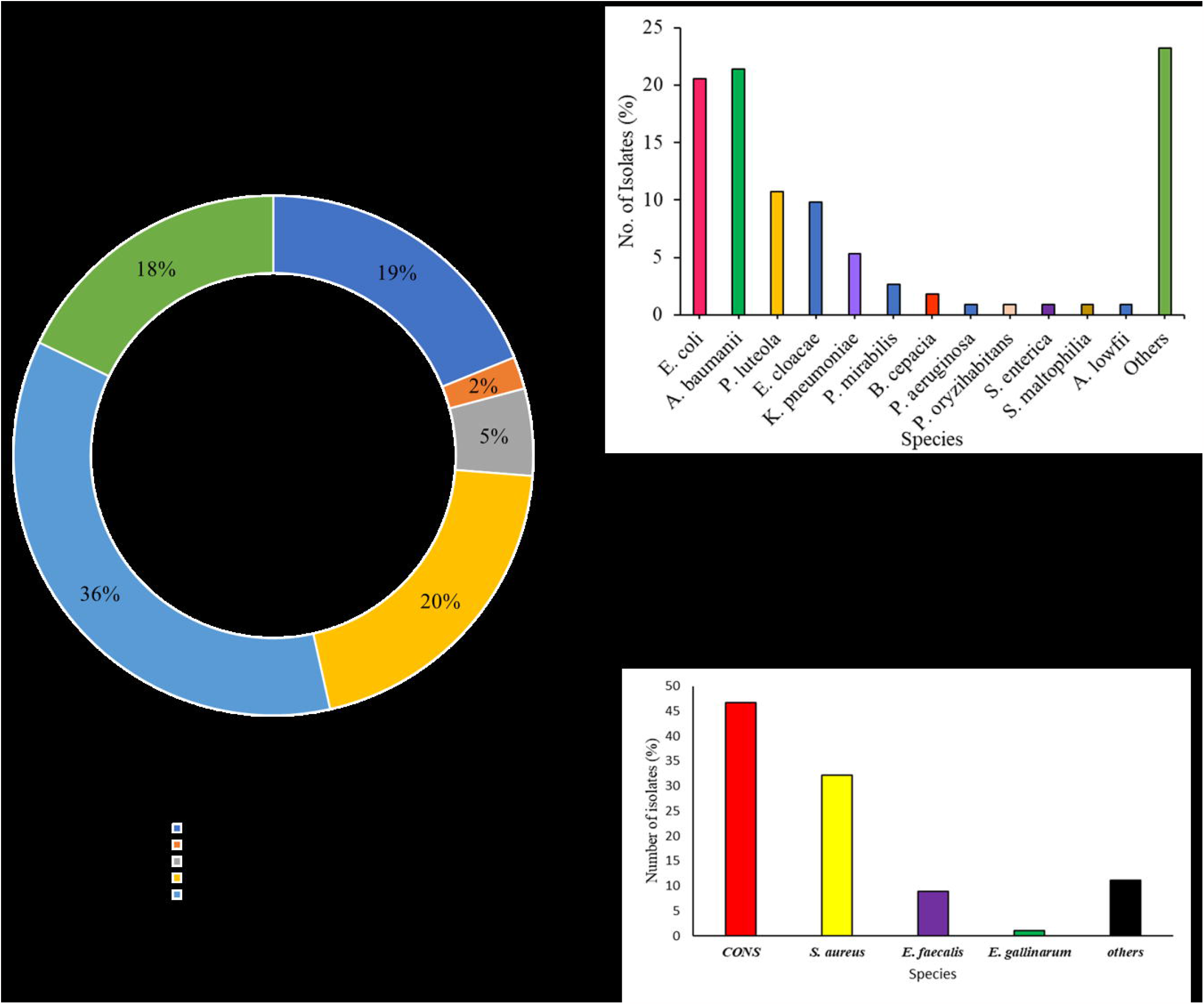
a) General distribution of bacterial isolates from patients and inanimate objects b)Spectrum of isolated gram-negative species; c) Isolated gram-positive species.

### Bacterial isolates from patients by sample types and wards

Coagulase negative *Staphylococci* (27/160, 17.5%) were the most isolated species followed by *S. aureus* (25/160, 15.6%) and *E. coli* (22/160, 13.8%) from clinical samples. The clinical sample with highest number of pathogens was pus swabs (92/160, 57.55%), followed by urine samples (56/160, 35%), stool (11/160, 6.87%) and lastly was the exit site swab (1/160, 0.7%). *E. coli, K. pneumoniae, K. oxytoca, P. mirabilis, S. enterica, E. cloacae and A. baumannii, CONs, S. aureus* and *E. faecalis* were all significant isolates from samples collected in this study. CONs and *S. aureus* are normal flora of the skin and normally contaminate wounds and urinary tract to cause infections in their unusual sites of habitation. The rate of isolation of other significant pathogens from the samples such as *E. cloacae* was (10/160, 6.25%) and *E. faecalis* (8/160, 5%). The most dominant isolates from urine were *E. coli* (12/56, 21.4%) followed by *A. baumannii* (7/56, 12.5%). *K. pneumoniae* were mainly from stool and pus swabs. *E. cloacae* and *E. feacalis* mainly found in pus 4/92 4.34% and urine samples (4/56, 7.14%) **Table 2**. *A. baumannii* (16.7%, 12/72) was highest in number amongst isolates from surgical ward **(Figure 2a)**. *S. aureus* (27.8%, 10/36) was the highest among all medical ward isolates **(Figure 2b)**. Among the clinical ICU isolates identified, *A. baumannii* was the most common at 36.3% (5/11) of all isolates in this section (**Figure 2c)**. The clinical renal unit isolates depicted that *E. coli* was the highest in number amongst all isolates with a percentage of 43.9% (18/41) (**Figure 2d)**.

**Table 2:**
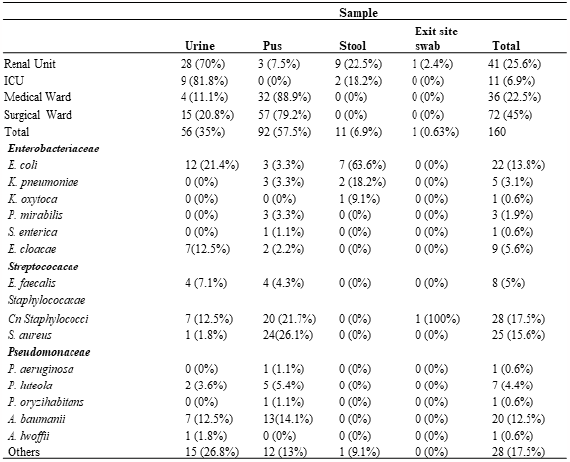
Distribution of identified species from different sample types by the site

**Figure 2.**
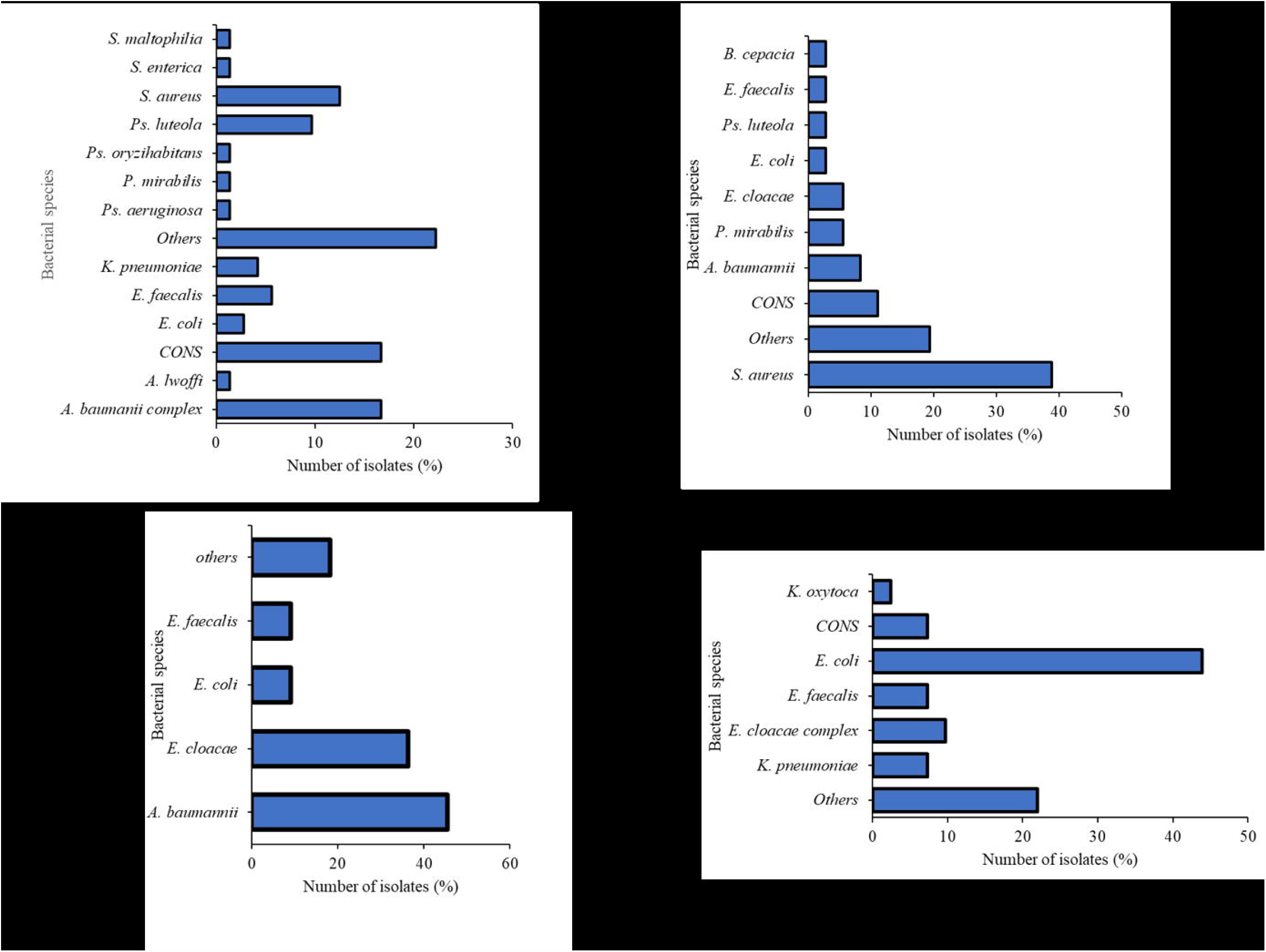
Isolates obtained from patients (A) shows isolates from surgical ward patients; (B) shows isolates from medical ward patients isolates; (C) shows isolates obtained from ICU patients and (D) shows isolates from renal unit patients.

### Distribution of bacterial isolates from patients by cases

Majority of bacterial isolates were from trauma patients in the surgical ward (67/160, 41.8%). The other bacterial isolates were distributed among burns patients 14/160 (8.75%), peritonitis patients 5/160, (3.1%), septic wound patients 3/160 (1.9%), decubitus ulcers patients 8/160 (5%) and renal unit patients with end stage renal disease (ESRD) 42/160 (26.23%), perforated gastric ulcer 4/160 (2.5%), degloving wound, cellulitis and others **(Figure 3)**.

**Figure 3.**
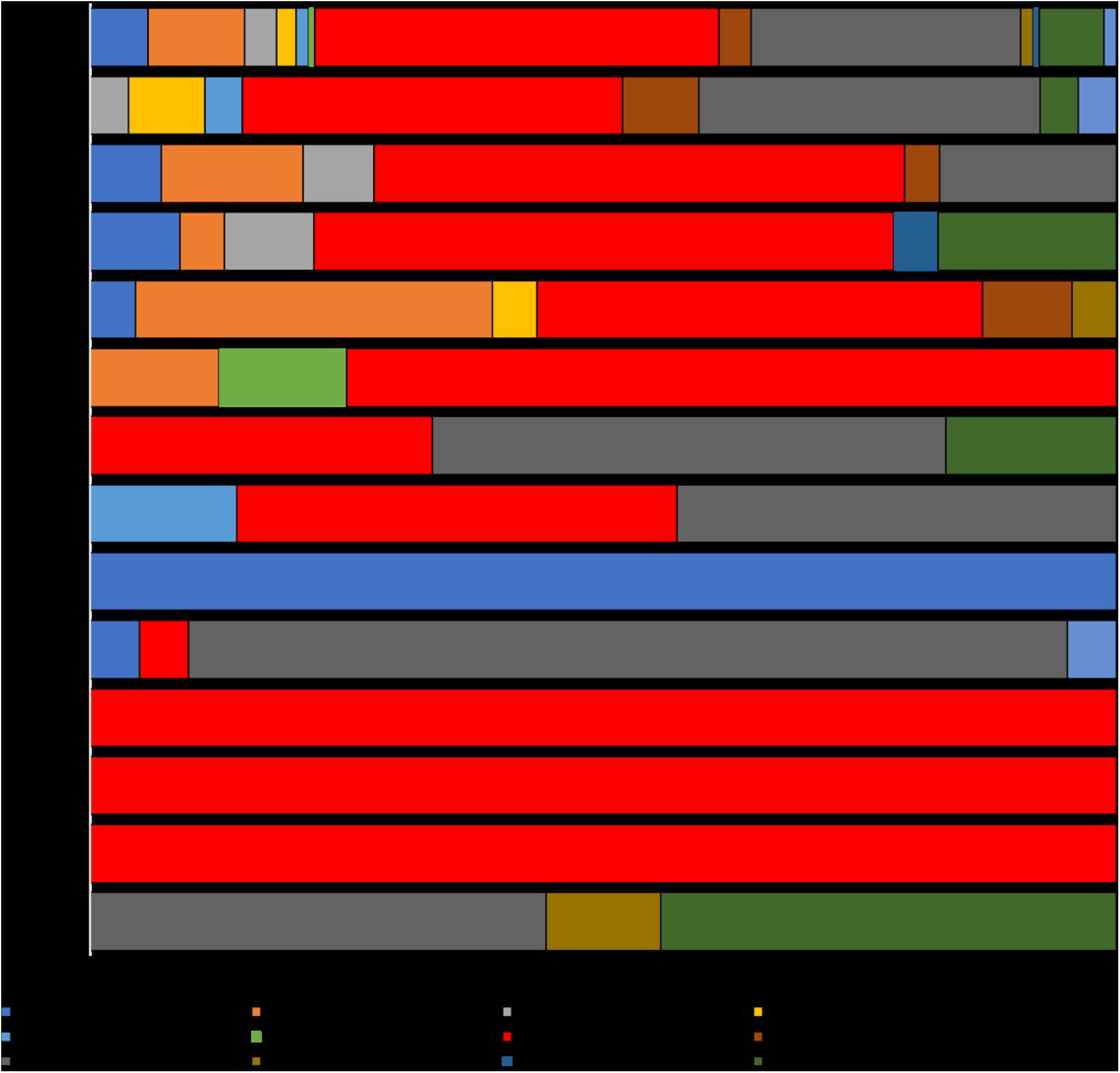
Distribution of various bacteria species isolated by patients’ presentations

### Bacterial isolates from hospital environment surfaces and inanimate objects

The bed rail was the most contaminated inanimate object sampled at 38% (16/42 isolates). This was followed closely by the monitors, which had 8/42 (19.0%) isolates, and the cabinets (7/42 isolates, 16.6%), of which both were placed in the ICU and renal units **(Figure 4a)**. Overall, coagulase negative *Staphylococci* isolates (CONs) were the most common bacteria isolated from high contact surface (38%, 16/42). This was followed by *S. aureus* (14.3%, 6/42) and *P. luteola* (9.5%, 4/42) (**Figure 4b**) all representing environmental swabs cultured from renal unit and ICU. The environmental isolates from ICU had *S. aureus* as the most common bacteria at 15.8% (6/38). In renal unit, four isolates were evenly distributed in the environment, namely, *S. epidermidis, S. lentus, E. cloacae* and *Y. pseudotuberculosis*.

**Figure 4.**
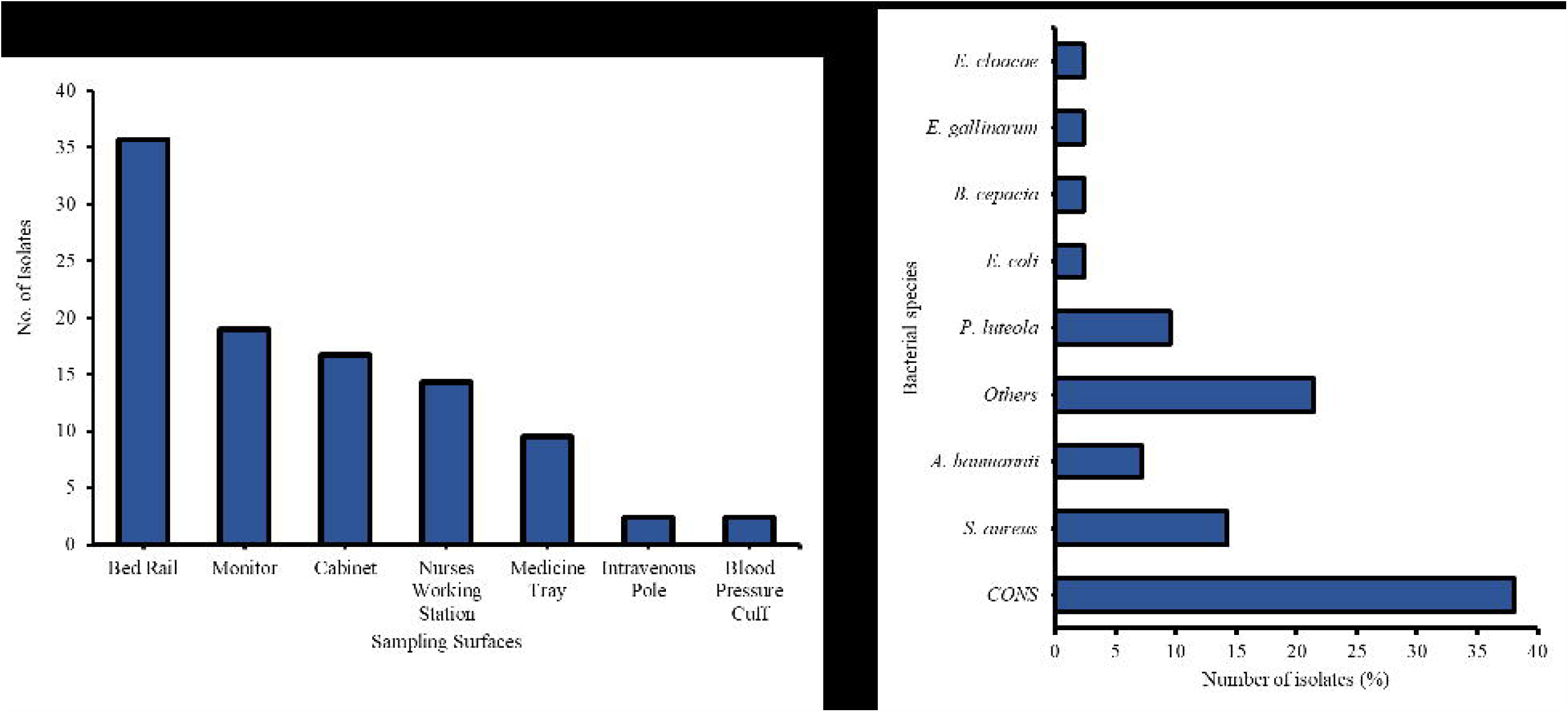
Bacterial isolates from hospital environment (A) Distribution on high touch surfaces (B) Type of isolates from high touch surfaces

### Antimicrobial susceptibility profiles of enteric bacteria and ESBL phenotype

AMR phenotypes of interest yielded included 29 isolates with ESBL phenotype which we characterized in this paper. Additionally, we characterized *A. baumannii, S. maltophilia* and *B. cepacia* resistance profiles. Overall, high resistance rates were observed against the third generation cephalosporins, monobactams, nalidixic acid, tetracyclines and the penicillins. Between 75-87% of the microorganisms were resistant to cephalosporins and ampicillin. Moderate level of resistance was observed against fourth generation cephalosporins (cefepime), amoxicillin clavulanate, piptazobactam, ciprofloxacin, sulphamethoxazole and gentamycin (50-65%) of the microorganisms. Low levels of resistance were observed against amikacin, minocycline, meropenem and imipenem (22-45% of the microorganisms). It is worth noting that the *E. cloacae* isolates were exceptionally highly resistant to those drugs that other enterics were moderately resistant against eg cefepime **(Figure 5a)**.

**Figure 5:**
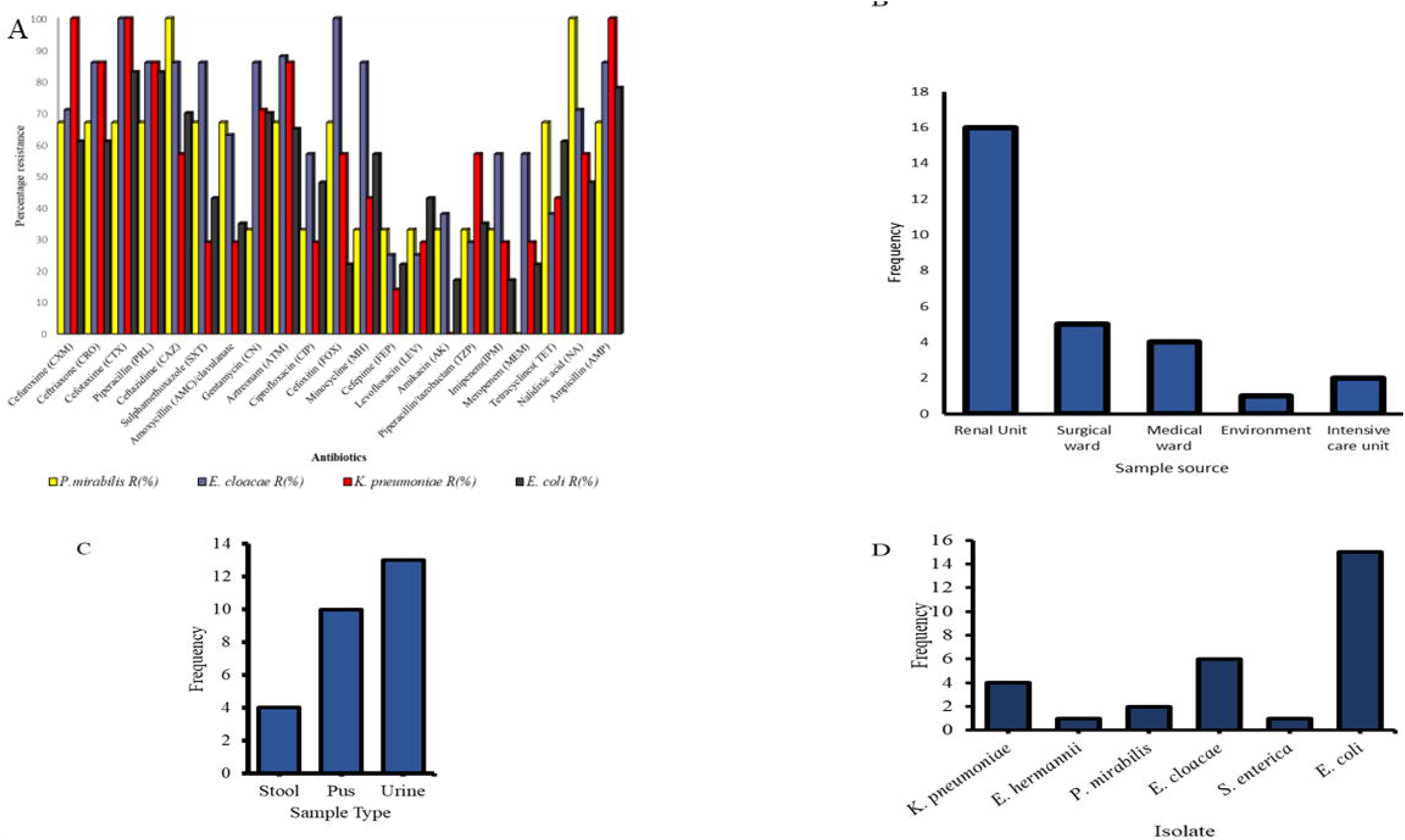
(A)Antibiotic resistance profiles of Enteric bacteria isolates (B) Distribution of ESBL phenotype isolates from different sections and wards (C)Distribution of samples from which bacteria portraying ESBL phenotype were isolated; (D) Frequency of enteric isolates with ESBL phenotype

Cefotaxime and piperacillin resistance among *E. coli* isolates was the highest at 83% 19/23. Ampicillin resistance rate came in second at 18/23 78%. Sulphamethoxazole rates of resistance followed at 70% (16/23). Ceftazidime (15/23) and aztreonam (15/23) resistance rates were both at 65.2%. Amikacin, cefoxitin, meropenem and imipenem remained highly effective against the *E. coli* isolates with resistance rates of 17% (4/23), 22% (5/23), 22% (5/23), and 17% (4/23) respectively **(Figure 5a)**. All the *K. pneumoniae* isolates in this study remained highly resistant against cefuroxime and cefotaxime at a rate of 100% (6/6). Ceftriaxone, piperacillin and ceftazidime resistant rates were also high at 83% (5/6). All *K. pneumoniae* isolates were susceptible to amikacin. Minocycline, ciprofloxacin, amoxicillin/clavulanate, imipenem and meropenem were highly effective against the *K. pneumoniae* isolates with resistance levels of (1/6)17%, 2/6, 33.2%, 2/6, 33.2% and 2/6 33.2% respectively **(Figure 5a)**. Statistical analysis showed that there was no significant difference in resistance rates across the different enteric bacteria p>0.05.

All the *Proteus mirabilis* isolates in this study remained highly resistant against ceftazidime (3/3) and nalidixic acid (3/3). Resistance against cefuroxime, ceftriaxone, cefotaxime and piperacillin came in second amongst *P. mirabilis* isolates at a rate of 67% (2/3). None of the *P. mirabilis* isolates was resistant to meropenem (0/3) and the resistance rate to imepenem was at 33 % (1/3) (**Figure 5a**). All the *Enterobacter* cloacae isolates in this study remained highly resistant against cefotaxime, cefoxitin at a rate of 100% (8/8). Ceftriaxone, piperacillin, amoxicillin and aztreonam were highly resistant as well at a rate of 87% (7/8). Levofloxacin and minocycline were the most effective drugs against *E. cloacae* with a resistance rate of 25% (2/8) (**Figure 5a**). The only *S. enterica* isolate from this study was resistant to all cephalosporins tested, monobactam, piperacillin, sulphamethoxazole, ampicillin, tazobactam-piperacillin, amoxicillin clavulanate, imipenem and meropenem.

A microorganism is presumed to be an ESBL producer when the diameters of zones of inhibition are below 25 mm for ceftriaxone, below 27 mm for cefotaxime, below 27mm for aztreonam and below 22mm for ceftazidime. 29/44 65.9% isolates were thus presumed to have the ESBL phenotype. In this study, renal unit isolates had the highest number of isolates with ESBL phenotype, followed by surgical ward and the medical ward **Figure 5b**. Urine samples from this study had the highest number of bacteria with ESBL phenotype followed by pus and stool samples **(Figure 5c)**. Majority of *E. coli* isolates (15/44, 34%) exhibited the ESBL phenotype, followed by *E. cloacae* (6/44, 13.6%) and *K. pneumoniae* (4/44, 9.09%) isolates (**Figure 5d)**.

The isolates seen in (**Figure 5d)** and **Table 3** were not only resistance to third generation cephalosporins and aztreonam but to many other antimicrobials such as tetracyclines and flouroquinolones. Most of these strains were susceptible to amikacin.

**Table 3:** Enteric bacteria and the antimicrobials they were resistant against

| Isolate | Source of isolate | Number of isolates (N) | Antimicrobials ineffective to enteric bacteria |
| --- | --- | --- | --- |
| *K. pneumoniae* | Pus | 3 | CXM, CRO, CTX, PRL, SXT, AMC, ATM, CAZ, FOX, FEP, TZP, NA, AMP |
| *E. hermanii* | Stool | 1 | CXM, CRO, CTX, PRL, ATM, CAZ, CIP, FEP, LEV, TZP, NA, AMP |
| *P. mirabilis* | Urine | 1 | CXM, CRO, CTX, PRL, AMC, ATM, CAZ, FOX, IMP, AMP |
| *E. cloacae* | Pus | 2 | CXM, CRO, CTX, PRL, SXT, AMC, CN, CAZ, FOX, TET, NA, AMP |
|  | Urine | 5 | CXM, CRO, CTX, PRL, SXT, AMC, CN, ATM, CAZ, CIP, FOX, FEP, NA, AMP |
| *E. coli* | Pus | 1 | CXM, CRO, CTX, PRL, SXT, AMC, CN, ATM, CAZ, CIP, FOX, FEP, LEV, TET, NA, AMP |
|  | Urine | 7 | CXM, CRO, CTX, PRL, ATM, CAZ, CIP, FEP, LEV, NA, AMP |
|  | Stool | 3 | CXM, CRO, CTX, PRL, SXT, AMC, CN, CAZ, CIP, FEP, MH, LEV, TZP, TET, NA, AMP |
| *S. enterica* | Pus | 3 | CXM, CRO, CTX, PRL, SXT, AMC, ATM, CAZ, CIP, FOX, FEP, LEV, TZP, TET, NA, AMP |
|  | Pus | 1 | CXM, CRO, CTX, PRL, SXT, AMC, ATM, CAZ, FOX, FEP, TZP, AMP |
|  | Total | 27 |  |

### Antimicrobial susceptibility profiles of *Acinetobacter spp, B. cepacia* and *S. maltophilia*

*A. baumanii* isolates were highly resistant to almost all groups of drugs including third and fourth generation cephalosporins, monobactams, aminoglycosides and the penicillins. For example, resistance to cefuroxime and aztreonam was at a rate 100% (**Figure 6)**. Considerable resistance to carbapenems was observed for these isolates. A high resistance rate to β-lactam inhibitors was observed with 100% to amoxicillin clavulanate and 95% for piptazobactam. Low levels of resistance were however recorded against minocycline 32%, levofloxacin (47%) and amikacin (47%).

**Figure 6:**
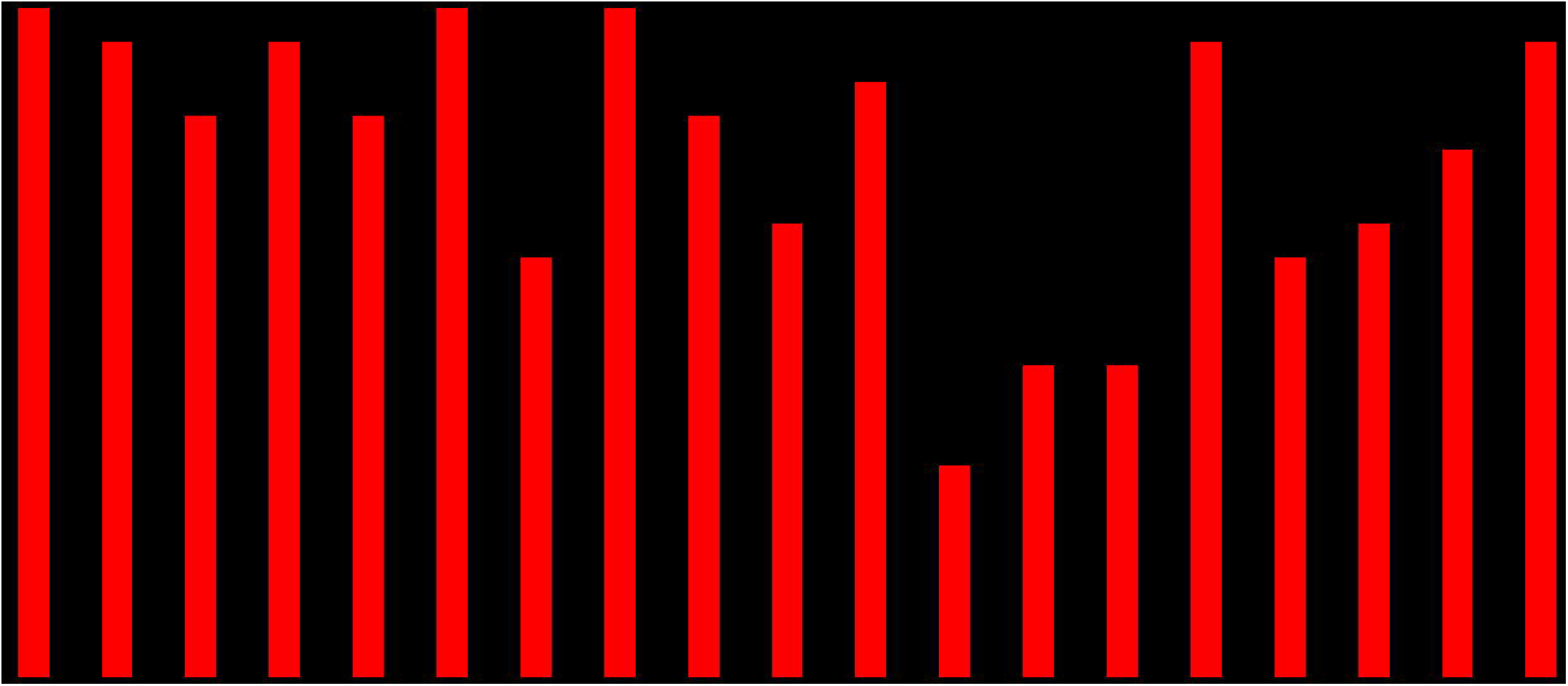
Resistance profiles of *A. baumannii*

*B. cepacia* and *S. maltophilia* isolates were resistant to almost all groups of drugs tested. Based on clinical laboratory standard institute (2020), interpretive zones for ceftazidime, meropenem, minocycline and sulphamethoxazole, all our *B. cepacia* isolates were found to be resistant against the four drugs **Table 4**. The *S. maltophilia* isolate was resistant to SXT and minocycline but was susceptible to levofloxacin **Table 4**.

**Table 4:** Antibiotic resistance results for *B. cepacia* and *S. maltophilia*

| Bacteria | Antibiotic agent | Result | Number |
| --- | --- | --- | --- |
| *B. cepacia* | Ceftazidime | R | 2/2 |
|  | Meropenem | R | 2/2 |
|  | Minocycline | R | 2/2 |
|  | Sulphamethoxazole | R | 2/2 |
| *S. maltophilia* | Sulphamethoxazole | R | 1/1 |
|  | Levofloxacin | S | 1/1 |
|  | Minocycline | I | 1/1 |

## Discussion

Bacterial infections contribute to patient’s poor prognosis, increased risk of ICU admission and mortality (4). The present study reveals a culture positivity rate that is higher than that of a study in Rwanda (19) and South Ethiopia (20). Among the positive samples, those associated with females were less than those from male’s contrary to other studies which demonstrated more samples from females than males (20). This was mainly due to the sharp increase in motor cycle accidents in Kenya, which mainly affects males (21). Gram-negative bacteria were the more dominant isolates similar to other studies conducted in Africa (20) (22). Overall, coagulase negative *Staphylococci* were the most isolated bacterial isolates, comparable to other studies that indicate CONS as the most commonly isolated microorganisms in clinical settings (19).

The bacterial spectrum was atypical from other studies (24,25)but here we describe the *S. maltophilia* and *B. cepacia* for the first time in our study site. *S. maltophilia* is a multidrug gram-negative bacillus that is an opportunistic pathogen, particularly for hospitalized patients (26) while *B. cepacia* is resistant to multiple antimicrobials and its treatment poses a challenge (27).

In the present study, most of the positive samples from the environmental were from ICU and constituted *S. aureus*. Similar results have been reported Ethiopia (28). The low number of positives in renal unit may be attributable to frequent fumigation that is done at the renal unit, done specifically after the dialysis sessions and effective infection prevention control mechanisms. These findings are echoed by other studies, which have reported no growth from dialysis ward bacterial surface culture (29).

Among the different surfaces and inanimate objects examined, the highest bacterial contaminated samples were taken from bed rails, cabinets, and monitors, corroborating the findings from similar studies that identified beds as highly contaminated (30) (31) (32). Overall, CONS were the most frequently isolated bacteria followed by *S. aureus*, consistent with findings from different studies from Ethiopia and elsewhere (31) (33). As for *P. luteola*, it made up 9.5% of isolates. The rate of isolation is comparable to other similar studies (34). Trauma patient samples yielded the largest number of bacterial isolates (33.1%) since most of them had undergone orthopaedic surgery and were in the surgical ward. This finding is similar to a previous study that recorded high rates of bacterial infection in orthopaedic patients (35).

In this study, *A. baumannii* was highest in number amongst isolates from surgical ward. Other studies have demonstrated that this species is frequently associated with skin and tissue infections at surgical sites (36) (37). Infections due to *S. aureus* was also high among all medical ward isolates.

In the present study, the sampling points for urine samples were ICU, surgical and medical wards which have long term admissions and catheterization. It has been reported that catheterization increases the incidence of urinary tract infection (UTI) by 3–5% and long admission increases the probability of acquiring UTI since hospitals are one of the sources of infection (38) Renal unit patients were revealed to be associated with the highest positive bacterial cultures of urine samples (50%) compared to all other clinical samples. This finding confirms that individuals with dysfunctional kidneys are more prone to urinary tract infections as previously reported (33).

Resistance to antimicrobial agents is a problem in health care facilities, but in hospitals, transmission of bacteria is amplified because of the highly susceptible population (39). The observed high resistance rates suggest poor adherence to antibiotic use guidelines and infection prevention policies in our study setting and beyond (40,41)The antibiotic sensitivity test results of our study confirmed that an alarming percentage of resistance were exhibited by bacterial isolates to the commonly prescribed antibiotics. Among the ESBL producers, the highest rates were observed amongst *E. coli* (32.6%) similar to other studies (42–44) and they were followed by *E. cloacae* isolates (18.6%). It has been reported that *E. cloacae* have selective advantage over other bacteria to produce Amp C when there is antibiotic pressure (45). The *E. cloacae* in this study demonstrated 100% resistance to cefotaxime and cefoxitin. Resistance to cephalosporins is mostly pronounced in these species (38). Enterobacteria from this study remained highly resistant to cefuroxime, cefotaxime, piperacillin and cefoxitin in general similar to previous studies (46). On the other hand, meropenem, imipenem, levofloxacin, and minocycline remained highly effective amongst the enteric bacteria.

*K. pneumoniae* isolates in this study were also highly resistant against cephalosporins but susceptible to minocycline, meropenem and imipenem. This pattern was also noted for *P. mirabilis* isolates. *E. coli* isolates highest resistant rates were against cefotaxime and piperacillin. Resistance rates for sulphamethoxazole was at 71% while that of ceftazidime and aztreonam were at 67%. The effective drugs against *E. coli* were amikacin and carbapenems. This is in consistency with a similar study carried out elsewhere (35). *Acinetobacter baumannii, Burkhholderia cepacia* and *S. maltophilia* isolates were highly resistant to all groups of drugs and this is in consistency with findings from other studies (47)(48)

## Conclusions

This study recorded presence of multidrug resistant organisms of concern including recently emerged ones such as *S. maltophilia* and *B. cepacia* at Thika Level V Hospital. Strategies to limit the spread of resistant strains should include encouraging the judicious use of antimicrobial agents. Hospitals can best reduce bacterial loads by improving waste-handling protocols, cleaning high-touch surfaces five times a day and providing soap at the handwash stations. Periodic monitoring systems should also be put in place to avert this problem. HAI control should be directed at the hospital and strengthened with different staff, supplies, budget, etc.

## Data Availability

All data prodcued in the present study are available upon reasonable request to the authors

## Acknowledgements

I would like to acknowledge help accorded by Gabriel Gikonyo during sampling in the hospital and Lydia Kwamboka who assisted in sample collection and culturing.

## Author contributions

RW, PM, MK, MM, WM, CA, BN and JG: Participated in the conception and design of the study, analysis and interpretations of the findings and drafting the manuscript. All authors reviewed and approved the final manuscript.

